# Human-specific evolutionary markers linked to foetal neurodevelopment modulate brain surface area in schizophrenia

**DOI:** 10.1101/2023.03.01.23286609

**Authors:** Maria Guardiola-Ripoll, Carmen Almodóvar-Payá, Angelo Arias-Magnasco, Mariona Latorre-Guardia, Sergi Papiol, Erick J Canales-Rodríguez, María Ángeles García-León, Paola Fuentes-Claramonte, Josep Salavert, Josep Tristany, Llanos Torres, Elena Rodríguez-Cano, Raymond Salvador, Edith Pomarol-Clotet, Mar Fatjó-Vilas

## Abstract

Schizophrenia (SZ) is hypothesised to represent a costly trade-off in the evolution of the neurodevelopmental ontogenetic mechanisms associated with human-specific cognitive capacities. Human Accelerated Regions (HARs) are evolutionary conserved genomic regions that have accumulated human-specific sequence changes. These evolutionary markers function as neurodevelopmental transcription enhancers and have been associated with the brain’s cortical expansion and connectivity, the processing of neural information, and the risk for SZ. We sought to investigate whether HARs’ polygenic load influenced neuroanatomical measures.

Our sample consisted of 128 patients with SZ and 115 healthy controls with high-resolution structural T1 MRI and genome-wide genotyping data. We extracted the cortical thickness (CT) and surface area (SA) for the 34 Desikan-Killiany regions per hemisphere. We calculated four polygenic risk scores (PRS): SZ genetic load (Global PRS_SZ_), HARs’ specific variability (HARs PRS_SZ_), HARs’ variability associated with transcriptional regulatory elements uniquely active in the foetal brain (FB-HARs PRS_SZ_) and in the adult brain (AB-HARs PRS_SZ_). Through linear regression analyses, we explored whether these four PRSs modulated CT and SA within diagnostic groups and the PRSs and diagnostic interaction on the cortical measures.

Results indicate that FB-HARs PRS_SZ_ influenced patients’ right SA on the lateral orbitofrontal cortex, the superior temporal cortex, the pars triangularis and the paracentral lobule and that a higher SZ risk load in FB-HARs is associated with lower SA values.

These findings evidence the involvement of the HARs-foetal gene regulatory activity in SA architecture and the evolutionary component of this regulation in SZ. These data emphasise the importance of HARs in the transcriptional regulatory machinery from early neurodevelopment and their role as the bridging point between the neurodevelopmental and evolutionary hypotheses in SZ.

## 1. INTRODUCTION

Schizophrenia (SZ) is a complex neuropsychiatric disorder characterised by symptoms that alter the perception and behaviour, such as hallucinations and delusions, and affectations of higher-order cognitive functions. These symptoms intimately relate the disorder with traits that distinguish humans as a species: abstraction, social cognition, language and thinking (Polimeni & Reiss, 2003). Accordingly, while the foundations of this multifactorial and complex disorder are not entirely understood, multiple pieces of evidence straightforwardly point towards a neurodevelopmental and evolutionary origin.

On the one hand, the strong genetic background of SZ, with heritability estimates up to 80% and a polygenic architecture with thousands of genetic variants with additive effects (Hilker et al., 2018; Legge et al., 2021; Purcell et al., 2009; Sullivan et al., 2003), converge with recent molecular data showing the role of developmental, neuronal, and synaptic differentiation pathways in the aetiology of the disorder (Gulsuner et al., 2013; O’dushlaine et al., 2015; Trubetskoy et al., 2022). The neurodevelopment processes and the neuroplasticity of the human brain are tightly orchestrated and involve gene expression regulatory mechanisms that are of paramount importance (Davidson, 2006). In line with the neurodevelopmental hypothesis of SZ, among the different environmental factors associated with an increased risk, prenatal and early adverse events occurring during crucial periods of brain development have been highlighted from epidemiological and clinical population-based studies due to their higher prevalence among people who later develop SZ (Rapoport et al., 2005). Also, the presence of prenatal complications was correlated with a higher genomic risk for the disorder (Ursini et al., 2018), and the placenta-associated genomic load has been linked to reduced brain volumes in neonates and poorer cognitive development during the first two years of life, emphasising the importance of the prenatal period in neurodevelopmental paths of risk (Ursini et al., 2021). In this line, delayed developmental milestones, considered the reflection of early deviances in neurodevelopmental trajectories, are associated with the disorder and predict psychotic symptoms in childhood and adulthood (Cannon et al., 2002; Niemi et al., 2003; Sørensen et al., 2010). Another aspect to be taken into account, is that prior to the onset of the psychotic symptoms, patients with SZ already present brain structural differences, such as reduction of grey matter, disrupted white matter integrity, and widespread cortical thinning (Haijma et al., 2013; Harrison et al., 2003), which latter remain once the diagnosis is settled (Jauhar et al., 2022; van Erp et al., 2018). Among the neuroanatomical measures used to characterise brain cortical morphology, cortical thickness (CT) and cortical surface area (SA) are highly heritable and influenced by largely independent genetic factors related to adult and mid-foetal active regulatory elements, respectively, but both related to neurodevelopmental genetic control (Grasby et al., 2020; Panizzon et al., 2009; Strike et al., 2019). All this evidence sustains the prevailing hypothesis that SZ results from environmental and genetic interactions modulating and deviating neurodevelopmental trajectories during the intrauterine and perinatal periods as well as during childhood and early adolescence (Birnbaum & Weinberger, 2017; Kahn et al., 2015) that disrupt the ontogenetic plan guiding brain architecture, brain configuration, and brain functioning.

On the other hand, the common prevalence of the disorder (nearly 1% (Charlson et al., 2018; J. McGrath et al., 2008; Perälä et al., 2007), and the fact that people affected, particularly males, have a reduced rate of reproduction (fitness) compared with the non-affected population (Haukka et al., 2003; J. J. McGrath et al., 1999) raise a question on why the genetic variants that increase the likelihood of suffering from SZ have persisted in the human genome? These, together with the close relationship between several clinical aspects of the disorder and human-specific cognitive traits (Polimeni & Reiss, 2003), have boosted the evolutionary view of the disorder. Accordingly, the evolutionary hypothesis of SZ suggests that the disorder emerged as a costly trade-off in the evolution of the ontogenetic mechanisms guiding human-specific neurodevelopment and sustaining complex cognitive abilities (Burns, 2004, 2006; Crow, 2000; Wynn & Coolidge, 2011).

While the evolutionary traces of SZ are difficult to follow, comparative genomics may be advantageous. The study of human-specific genomic changes may lead to a better comprehension of human-specific phenotypic traits and increased knowledge of what genetic changes contributed to making us human (O’Bleness et al., 2012). In this sense, Human accelerated regions (HARs) might be helpful. HARs are evolutionary conserved genomic regions that have experienced significant changes after human and chimpanzee divergence (Bird et al., 2007; Bush & Lahn, 2008; Gittelman et al., 2015; Lindblad-Toh et al., 2011; Pollard, Salama, King, et al., 2006; Prabhakar et al., 2006). This accelerated divergence of HARs is suggested to reflect their role in some human-specific characteristics. Most HARs are intergenic, within introns near protein-coding genes, transcription factors and DNA binding proteins (Capra et al., 2013; Doan et al., 2016; Hubisz & Pollard, 2014; Pollard, Salama, Lambert, et al., 2006; Won et al., 2019). All the studies that intended to characterise HARs’ functional role converge in highlighting them as transcription factors binding sites, transcription factors on their own and participants in the neurodevelopmental gene expression machinery (Doan et al., 2016; Girskis et al., 2021; Uebbing et al., 2021; Won et al., 2019).

Recently, studies inspecting the expression patterns of HARs-associated genes (HARs-genes) show their implication in human-specific cortical expansion, brain functional connectivity and the brain’s neural information processing. First, in a comparative study exploring the cortical expansion in humans and chimpanzees, it was described that the expression profiles of HARs-genes correlated with the expansion of higher-order cognitive networks, such as the frontoparietal and the default mode networks (Wei et al., 2019). The same study also revealed that the genetic variability in HARs-genes expressed in the brain (HARs-brain genes) was associated with the DMN functional variation in healthy subjects (Wei et al., 2019). Second, HARs-brain genes expression patterns have been related to the individual variability in functional connectivity and to the brain’s information processing (Li et al., 2021; Luppi et al., 2022). Remarkably, these studies report that HARs-brain genes show the highest expression in higher-order cognitive networks, such as the frontoparietal and the default mode networks; the ones with the greatest functional heterogeneity across individuals and also the ones with predominant synergistic interactions.

The evidence on HARs’ contribution to human-specific brain architectural configuration, functioning and information processing is also accompanied by studies that describe that HARs’ genetic variability influences the risk for SZ. For example, the investigation into the overlap between HARs and SZ GWAS SNPs showed that SZ’s polygenic background was enriched in genes associated with these evolutionary regions (Xu et al., 2015). In line, subsequent findings also described that SNPs in HARs or in linkage disequilibrium with them were more likely associated with the disorder (Srinivasan et al., 2017). Notwithstanding, to our knowledge, HARs modulation of brain measures in SZ has been scarcely explored, and further studies using brain-based phenotypes to assess their role in the disorder are necessary.

Considering the polygenic nature of both cortical structural configuration and SZ’s susceptibility, studies using measures summarising this complex genetic architecture, such as Polygenic Risk Scores (PRSs), would be helpful to disentangle the genetic roots not only of the disorder but of complex brain traits. The PRS is a quantitative measure of the genetic burden of a trait based on GWAS data. It can be calculated at a whole-genome level, but also within subsets of SNPs defined based on their involvement in particular biological pathways of interest. Therefore, based on the evidence of HARs’ role in neurodevelopment, brain configuration and susceptibility for SZ, we aimed to investigate the modulatory effect HARs’ polygenic load on neuroanatomical measures through a neuroimaging genetics approach in healthy control and patients with SZ. We generated different PRSs summarising HARs genetic variability, specifically including HARs SNPs related to active regulatory elements in the foetal and adult brain. We explored whether the PRSs modulated cortical thickness and surface area differently depending on the health/disease condition.

## 2. MATERIAL AND METHODS

### Sample

The initial sample consisted of a case-control dataset of 378 individuals, of which 284 passed both the genetic and neuroimaging quality control (see details in the corresponding *Molecular analyses* and *MRI data acquisition* sections). Patients were recruited from the inpatients and outpatients at the Hospital Benito Menni, Sant Boi de Llobregat (Barcelona, Spain) and healthy controls were recruited from the same area. The patients’ diagnosis was confirmed according to DSM-IV-TR based on an interview with two psychiatrists. All participants were of European ancestry, between 18 and 65 years old, right-handed and had an estimated intelligence quotient (IQ) (premorbid IQ in patients), higher than 70, as assessed using the Word Accentuation Test (Gomar et al., 2011). All participants met the same exclusion criteria, which included suffering from major medical illness, conditions affecting cognitive or brain function, neurological conditions, history of head trauma with loss of consciousness and present or history of drug abuse or dependence. Additionally, for healthy controls (HC), exclusion criteria also included personal or family history of psychiatric service contact or treatment.

Individuals in both diagnostic groups were matched by age and sex to minimise age and sex differences between them, and the analyses were conducted in a sample of 115 HC and 128 patients with a SZ diagnosis (**Table 1**).

**Table 1.**
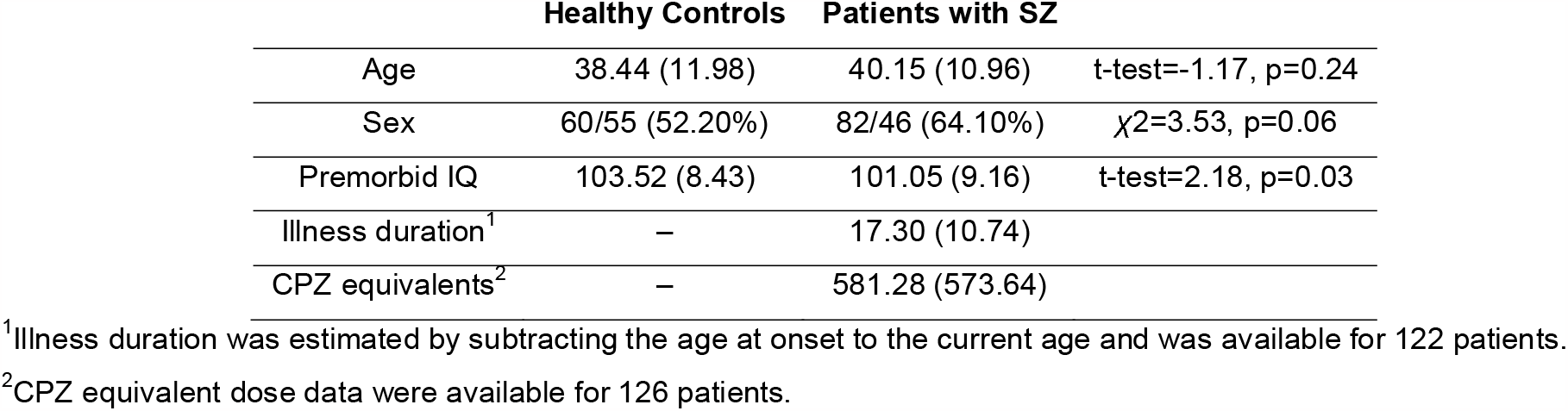
Sample characteristics, including demographic and clinical description. All the quantitative variables include mean and standard deviation (sd). Sex description includes male/female count (% of males). Illness duration is given in years and Chlorpromazine (CPZ) equivalents in mg/day.

All subjects signed a written consent after being fully informed about the procedures and implications of the study, approved by the Germanes Hospitalàries Research Ethics Committee, and performed following its guidelines and in accord with the Declaration of Helsinki.

### Molecular analyses

Genomic DNA was extracted either from buccal mucosa through cotton swabs using ATP Genomic Mini Kit Tissue (Teknokroma Analitica, S.A., Sant Cugat del Valles, Spain) or from peripheral blood cells using Realpure SSS kit (Durviz, S.L.U., Valencia, Spain).

A genome-wide genotyping was performed using the Infinium Global Screening Array-24 v1.0 (GSA) BeadChip (Illumina, Inc., San Diego California, U.S) at the Spanish National Cancer Research Centre, in the Human Genotyping lab (CeGen-ISCIII), resulting in the genotyping of 730,059 SNPs. After QC, a dataset of 447,035 SNPs with the following characteristics was obtained: Hardy-Weinberg equilibrium in patients and healthy controls, SNP call rate higher than 98% and minor allele frequency (MAF) higher than 0.005. Individuals with an SNP missingness higher than 2% were excluded. In addition, through a principal component analysis (PCA), those individuals found to be related or not of European ancestry were also excluded. Next, re-phasing and imputation were performed using, respectively, Eagle (Durbin, 2014) and Minimac4 (Das et al., 2016) and the Haplotype Reference Consortium dataset (HRC version r1.1) (McCarthy et al., 2016) hosted on the Michigan Imputation Server (Das et al., 2016) (https://imputationserver.sph.umich.edu/). A MAF value of > 1% and an imputation quality of R^2^ > 0.3 were required for the inclusion of the variants into further analyses. Finally, our final SNP dataset included 7,606,397 genetic markers.

### Polygenic Risk Score Estimation

Using SZ GWAS 2022 summary statistics from the European subsample (Trubetskoy et al., 2022) we estimated four different PRS using PLINK 1.90 software (Chang et al., 2015) based on the PRS-C+T methodology (Privé et al., 2019), in a similar fashion as several recent GWAS studies (Grasby et al., 2020; Mullins et al., 2021; Trubetskoy et al., 2022). This method is defined as the sum of allele counts, weighted by estimated effect sizes obtained from the GWAS, after two filtering steps: LD clumping (based on the European population from the phase 3 1,000 Genomes reference panel) and p-value thresholding.

First, we calculated the whole-genome PRS (**Global PRS**_**SZ**_). The LD filtering was conducted by including the most significant SNP from any pair showing an LD r^2^>0.015 within 1,000kb windows, resulting in a set of informative linkage-disequilibrium independent markers (98,121 SNPs). Subsequently, for the p-value thresholding, we considered a range of thirteen p-value thresholds: p<5×10^−8^, p<5×10^−7^, p<5×10^−6^, p<5×10^−5^, p<5×10^−4^, p<5×10^−3^, p<0.05, p<0.1, p<0.2, p<0.3, p<0.4, p<0.5, p<1.0. Through logistic regression, we established the best threshold in p<5×10^−3^ as the better predictor of the diagnosis status () based on the Nagelkerke’s pseudo R^2^ (p=4.75×10^−12^, R^2^=0.22).

Second, we estimated a PRS accounting for HARs genetic variability (**HARs PRS**_**SZ**_), exclusively including SNPs within the 3,070 autosomal HARs sequences compiled by Girskis et al., 2021 (*Supplementary Table 1*). Using Bedtools 2.30.0 (Quinlan & Hall, 2010) we selected the genetic markers in our sample within these HARs sequences. Considering the number of SNPs within the HARs, the PRS estimation was conducted following the same PRS-C+T methodology but adjusting LD clumping parameters. Following PLINK default options, we selected the most significant SNPs within 250kb windows and with LD r^2^>0.5 and the unique p-value threshold was set at p<1.0. The final set of variants was composed of 2,114 SNPs.

Third, to assess the effect of HARs SNPs specifically affiliated with foetal brain (FB) or adult brain (AB) gene regulatory elements, we estimated two additional PRS scores (**FB-HARs PRS**_**SZ**_ and **AB-HARs PRS**_**SZ**_) with the same procedure and parameters as for PRS-HARs. Following the methodology used in the latest ENIGMA human cerebral cortex GWAS by (Grasby et al., 2020), we downloaded ChromHMM chromatin states (core 15 state model) from the Epigenomics Roadmap (Roadmap Epigenomics Consortium et al., 2015). We selected two foetal tissues (E081=foetal brain female and E082=foetal brain male) and four adult tissues (E067=brain angular gyrus, E069=brain cingulate gyrus, E072=brain inferior temporal lobe and E073=brain dorsolateral prefrontal cortex) and for each tissue, the genomic regions comprising active regulatory elements (TssA, TssAflnk Enh and EnhG). We combined the foetal (E081 and E082) and adult (E067, E069, E072 and E073) annotations and selected only those regions non-overlapping between them as foetal brain-specific and adult brain-specific. With the selected **HARs PRS**_**SZ**_ SNPs, we selected the genetic variants allocated within these foetal and adult brain-specific regions. The final set of variants included in the **FB-HARs PRS**_**SZ**_ and **AB-HARs PRS**_**SZ**_ estimations were 112 and 81 SNPs, respectively.

### MRI data acquisition

The MRI neuroimaging data were obtained from two scanners: 58% (70 HC, 72 patients) of the sample was scanned in a 1.5T GE Sigma scanner (General Electrical Medical Systems, Milwaukee, Wisconsin, USA) and the other 42% (45 HC, 56 patients) in a 3T Philips Ingenia scanner (Philips Medical Systems, Best, The Netherlands) at Hospital Sant Joan de Déu (Barcelona, Spain).

High-resolution structural-T1 MRI data in the 1.5T scanner was obtained using the following acquisition parameters: matrix size 512 × 512; 180 contiguous axial slices; voxel resolution 0.47 × 0.47 × 1 mm^3^; echo time (TE) = 3.93 ms, repetition time (TR) = 2,000 ms; and flip angle = 15°. At the 3T scanner, structural T1-weighted sequences were acquired as follows: matrix size□320□×□320 × 250; voxel resolution 0.75 × 0.75 × 0.80 mm3; TE= 3.80 ms, TR = 8.40 ms; and flip angle = 8°. All images were visually inspected to exclude those with artefacts and movement.

### Surface-based morphometry

Structural MRI data were processed using the FreeSurfer image analysis suite (http://surfer.nmr.mgh.harvard.edu/). Image pre-processing included removal of non-brain tissue, automated Talairach transformation, tessellation of the grey and white matter boundaries and surface deformation (Fischl et al., 2004). Several deformation procedures were performed in the data analysis pipeline, including surface inflation and registration to a spherical atlas. This method uses both intensity and continuity information from the entire three-dimensional images in the segmentation and deformation procedures to produce vertex-wise representations of CT and SA. The CT was defined as the measure of the distance between the white matter surface and the pial surface, and cortical SA was calculated as the area of the white matter surface. With FreeSurfer we automatically performed the segmentation of 34 cortical regions of interest for each hemisphere using the Desikan-Killiany cortical atlas (Desikan et al., 2006). Mean values of CT and SA were quantified for each individual within these defined regions.

All subjects included in this study passed the standardised quality-control protocols from the ENIGMA consortium (https://enigma.ini.usc.edu/protocols/imaging-protocols/) that have previously been applied in large-scale multi-centre studies (Grasby et al., 2020; Hibar et al., 2018).

### Statistical analyses

Demographic and clinical data were processed and analysed using SPSS (IBM SPSS Statistics, version 29.0, released 2022, IBM Corporation, Armonk, New York, USA).

We compared the SZ polygenic load of patients and controls based on the four different PRS estimations (Global PRS_SZ_, HARs PRS_SZ_, FB-HARs PRS_SZ_ and AB-HARs PRS_SZ_) by means of block-wise logistic regressions with the two diagnostic groups with SPSS. For each PRS, two statistical models with case/control status as outcome were compared, one testing the covariates alone (age, sex and the two first ancestry-specific principal components as a baseline model), and the other testing the covariates plus the corresponding PRS (full model). We report the R^2^ values as the differences in Nagelkerke’s pseudo-R^2^ between these two nested models as an indicator of explained variance (Smigielski et al., 2021; Smith & Mckenna, 2013).

Next, we examined to which extent different PRSs explain CT and SA measures from the 34 cortical regions. We applied linear regression models (R software) within diagnostic groups (separately in HC and patients with SZ) to test the effect of each PRS on CT/SA. In order to assess whether the PRS effect was modulated by the diagnostic status, we conducted linear models using the whole sample and tested the PRS x diagnosis interaction. Sex, age, premorbid IQ, intracranial volume, and scanner effect were included in the statistical models in order to control for their potential effects. In the linear regression within patients with SZ, the antipsychotic dose was as well included as a covariate (Scherk & Falkai, 2006; van Erp et al., 2018).

According to the findings, the SNPs included in the FB-HARs PRS_SZ_ were furtherly mapped and functionally annotated using FUMA (Watanabe et al., 2017) with the *SNP2GENE* and the *GENE2FUNC* tools. The positional mapping parameters were left as default. The eQTL mapping was conducted in PsychENCODE, ComminMind and BRAINEAC tissues filtering by PsychiENCODE and brain open chromatin atlas annotations. The 3D chromatin interaction mapping was conducted with PsychENCODE, adult and foetal cortex, dorsolateral and hippocampus and neural progenitor cells data filtering by PsychENCODE and brain open chromatin atlas annotations.

The p-values resulting from each one of the before mentioned statistical tests were adjusted by using the false discovery rate (FDR) method, specifically the Benjamini-Hochberg procedure, to control for multiple comparisons at level q=0.05. Accordingly, only those results with a corrected FDR-pval<0.05 were considered statistically significant.

The significant results on the cortical regions were plotted using the *ggseg* library in R (Mowinckel & Vidal-Piñeiro, 2020) and the regressions with the direction of the results were plotted using SPSS (**Figure 1**).

**Figure 1.**
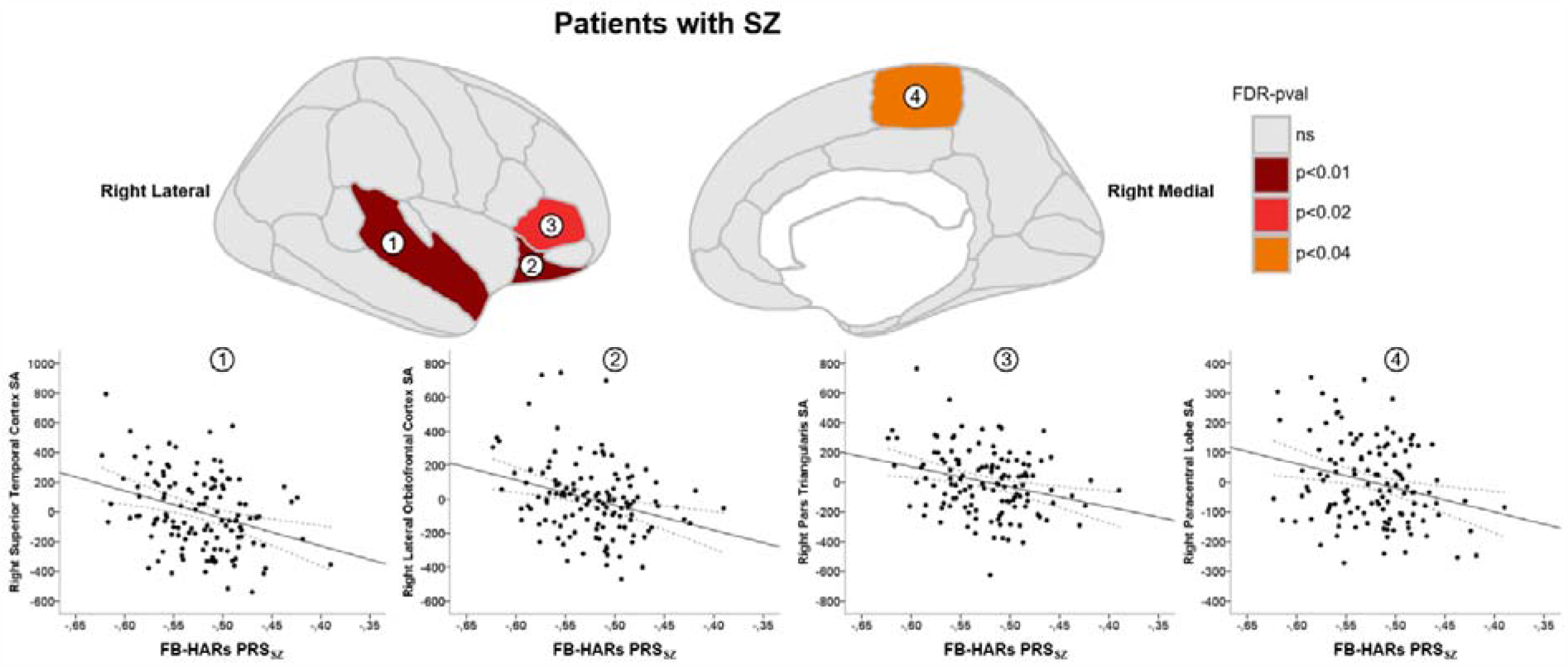
Brain and scatter plots with significant FB-HARs PRS_SCZ_ effect on surface area in patients with schizophrenia (SZ). Brain plots include the lateral and medial sagittal views for the right hemisphere. The coloured regions are the ones with significant FB-HARs PRS_SCZ_ effect on surface area (SA) after FDR correction (FDR-pval < 0.05). The scatter plots show the relationship between the FB-HARs PRS_SCZ_ (on the X-axis) and the unstandardised SA residuals (on the Y-axis in cm^2^, estimated regressing out the covariates) and evidence their negative correlation (black solid line and black dashed lines representing mean 95% confidence intervals). Each region is numerically labelled as following: 1) superior temporal cortex; 2) lateral orbitofrontal cortex; 3) pars triangularis; and 4) paracentral lobule.

## 3. RESULTS

### PRS comparison between diagnostic groups

Diagnostic PRS comparisons revealed differences in the Global PRS_SZ_, with patients presenting higher SZ genetic load than HC. The HARs-derived PRSs did not show between-groups differences (**Table 2**).

**Table 2.** Polygenic risk score (PRS) comparisons between healthy controls (HC) and patients with schizophrenia (SZ). PRS means and standard deviations (sd) are given for both diagnostic groups for the four estimated PRSs (Global PRSSZ, HARs PRSSZ, FB-HARs PRSSZ, AB-HARs PRSSZ). The logistic regression results include the β and standard error (se), the logistic regression statistic (Wald, W), the Nagelkerke’s pseudo R^2^ (R^2^), and the adjusted p-values after FDR correction (FDR-pval).

| | HC mean<br>(sd) | SCZ mean<br>(sd) | $\beta$ (se) | W | $R^2$ | FDR-pval |
| --- | --- | --- | --- | --- | --- | --- |
| **Global PRS<sub>SZ</sub>** | -201.14<br>(1.23) | -200.10<br>(1.02) | 0.79 (0.14) | 32.97 | 0.20 | $3.75 \times 10^{-8*}$ |
| **HARs PRS<sub>SZ</sub>** | -5.45 (0.17) | -5.41 (0.19) | 1.32 (0.73) | 3.28 | 0.02 | $9.33 \times 10^{-2}$ |
| **FB-HARs PRS<sub>SZ</sub>** | -0.53 (0.05) | -0.52 (0.04) | 2.07 (2.96) | 0.49 | 0.003 | $4.85 \times 10^{-1}$ |
| **AB-HARs PRS<sub>SZ</sub>** | -0.43 (0.04) | -0.42 (0.04) | 7.69 (3.60) | 4.55 | 0.02 | $6.57 \times 10^{-2}$ |

\*Significant findings at FDR-pval<0.001.

### PRS associations with morphometric measures

Regarding CT measures, no PRS estimates modulated thickness either within HC or patients with SZ. In the same line, the PRSs x diagnosis interactions on CT evidenced no significant effects.

In contrast, the linear regression analyses revealed that among patients with SZ, the FB-HARs PRS_SZ_ significantly affected SA in different regions in the right brain hemisphere. FB-HARs PRS_SZ_ modulated the patients’ SA of the lateral orbitofrontal cortex (Standardised β=-0.234, SE=440.443, adjusted R^2^=0.491, FDR-pval=0.008), the superior temporal cortex (Standardised β =-0.235, SE=545.898, adjusted R^2^=0.488, FDR-pval=0.008), the pars triangularis (Standardised β=-0.242, SE=438.928, adjusted R^2^=0.322, FDR-pval=0.019) and the paracentral lobule (Standardised β=-0.233, SE=282.910, adjusted R^2^=0.264, FDR-pval=0.030) (**Figure 1**). In these regions, a higher SZ risk load in FB-HARs was associated with lower SA values. Conversely, no other associations were observed between the other estimated PRSs and SA neither within HC nor in interaction with diagnosis.

The inspection of the genomic context of the SNPs in FB-HARs PRS_SZ_, showed that 54.1% of the SNPs were in intergenic regions, 26.5% in introns and 16.6% in intronic non-coding RNA. They mapped into 223 genes (*2*). The subsequent functional annotation results highlighted that these genes were enriched in several Gene Ontology (GO) categories. The biological process retrieved were neuron differentiation, neurogenesis, neuron development, and forebrain development, among others. Also, the only cellular component category enriched was cell junction (*Supplementary Table 3*).

## 4. DISCUSSION

Through a neuroimaging genetic approach, we have evaluated the SZ’s polygenic load of specific human evolutionary markers, such as HARs, on brain-based phenotypes closely related to the pathophysiology of the disorder. This is, to the best of our knowledge, the first study assessing the effect of HARs genetic variability on brain cortical measures in patients with SZ and healthy controls. Our analyses provide evidence on the modulatory effect of foetal active regulatory HARs on the cortical surface area of different brain regions in patients with SZ. These findings highlight the importance of human-specific genetic changes, especially those affecting active regulatory elements specific to foetal neurodevelopment, in the genetic machinery guiding human brain cortical structure.

The comparisons between the different PRS estimates across diagnostic groups show that individuals with a diagnosis of SZ present higher Global PRS_SZ_ than healthy individuals. Thus, the sample of patients presents a higher polygenic load for SZ. This result aligns with the current view on the value of the PRS_SZ_ as a highly informative genetic vulnerability marker for its consistency across numerous studies, not only in patients’ *vs* controls comparisons (Calafato et al., 2018; Vassos et al., 2017) but through family approaches, which evidence the intermediate genetic load that healthy relatives of affected patients have (Smigielski et al., 2021; van Os et al., 2020).

However, in our sample, the analyses indicate no significant differences across diagnostic groups using the other three HARs-related PRS_SZ_. The direct comparison of our findings with previous studies is difficult because of the absence of HARs-based PRS studies; nevertheless, some previous studies have explored the role of specific candidate HARs in SZ through association approaches. In this sense, the haplotypic variability at the *HAR1A* gene, a novel RNA gene with a presumable neurodevelopmental role that harbours the HAR with the highest substitution rate in humans as compared to chimpanzees (Pollard, Salama, Lambert, et al., 2006), was associated with auditory hallucinations in patients with a schizophrenia-spectrum disorder in a European sample (Tolosa et al., 2008). In line, several candidate HARs-SNPs altering transcription factor binding sites and presenting methylation marks of active promoters, repressors or enhancers in the brain were associated with the risk for SZ and modulated cognitive performance in a north-Indian population (Bhattacharyya et al., 2021, 2022). On the other hand, genome-wide-based studies also describe that HARs-genes and HARs-brain genes are associated with SZ at the GWAS level (Cheung et al., 2022; Srinivasan et al., 2017; Wei et al., 2019; Xu et al., 2015). Some of these studies, indeed, went beyond common variability and showed that rare variants in HARs-genes were also enriched with the disorder (Wei et al., 2019); suggesting, therefore, that common and rare variants joint analyses can help disentangling the role of HARs variability on the susceptibility for the disorder and its specific phenotypes.

In our investigation on the contribution of HARs polygenic background on cortical neuroanatomical measures variability, we report a modulatory effect of FB-HARs PRS_SZ_ on the SA within patients with SZ. These findings suggest that the genetic variability in HARs associated with regulatory elements uniquely active in the foetal brain would specifically influence brain phenotypes in SZ. Results show that as the FB-HARs PRS_SZ_ increases (i.e as more SZ risk variants accumulate in these HARs associated with foetal active regulatory elements), patients present lower cortical surface area in the lateral orbitofrontal cortex, the superior temporal cortex, the pars triangularis and the paracentral lobe. First, these findings converge with data highlighting the importance of the foetal period (and the gene expression circumscribed to it) to understand the neurodevelopmental trajectories linked to schizophrenia (Ursini et al., 2021). Also, our findings line up with previous studies reporting widespread smaller surface area, with the largest effect sizes in the frontal and temporal lobe regions (van Erp et al., 2018). Focusing on the regions significantly modulated by the SZ genetic load in foetal brain active HARs, we can draw attention to the orbitofrontal region and the temporal cortex as these are among the regions suffering the largest expansion in the human cortex in comparison with chimpanzees. Wei et al., 2019 described that areas of the orbital frontal gyrus and the temporal lobe experienced an x4 and x3 expansion, respectively, and evidenced that the transcription profile of 1711 HARs-genes positively correlated to the pattern of human cortical expansion, meaning that the highest HARs-gene expression occurs in highly expanded areas of the human cortex.

A remarkable finding though is the specificity of significant PRS association with SA, as we were not able to observe HARs polygenic effect on the CT variability. This could be interpreted considering that the genetic influences on the two cortical measures and the underlying mechanisms are largely different, and that SA and CT follow distinct developmental trajectories (Jha et al., 2018; Lyall et al., 2015; Wierenga et al., 2014). As posited by the radial unit hypothesis, cortical surface area expansion would be driven by the proliferation of neural progenitor cells, while thickness would be determined by the number of their neurogenic divisions (Rakic, 1988). In this line, a study described that HARs-genes are highly expressed during prenatal development, their expression is upregulated during neurogenesis and enriched in cells from the outer radial glia (Won et al., 2019). Indeed, radial glia is a major class of neural stem cells in the germinal layer that shows substantial expansion in the primate lineage and among the neurodevelopmental differences between human and chimpanzees there is the proliferative capacity of neural progenitors during cortical development (Mora-Bermúdez et al., 2016). Recent data shows that the genetic determinants of SA are predominately related to gene regulatory activity in neural progenitor cells during foetal development while CT is influenced by regulatory processes that occur after mid-foetal development (Grasby et al., 2020). Also, common genetic variants explained a larger part of SA variance (SNP-h^2^=34%, SE=3%) than of CT variance (SNP-h^2^=26%, SE=2%) (Grasby et al., 2020). However, our results could also be influenced by the reduced number of SNPs in our PRS estimates, which could have hampered the capture of the genetic determinants of CT. Likewise, by using HARs, we are putting the focus on regions highly stable along mammal evolution that experiences rapid sequence changes in the human lineage since the divergence from our closest relatives (Girskis et al., 2021), and while SA has enormously increased during the evolution of primates, cortical thickness has remained relatively constant (Eickhoff et al., 2005). Therefore, other genetic evolutionary markers could be more suitable for inspecting the evolutionary traces of CT.

Relative to the exploratory gene mapping results, it was interesting to find that the SNPs underlying cortical surface area differences within patients were enriched in biological processes important for nervous system development. These findings would be in line with previous studies describing that HARs associated genes mainly participate in biological processes and pathways related to neurodevelopment, neural differentiation and axonogenesis (Doan et al. 2016).

Finally, we should account for some limitations of this study. Regarding our genetic association approach, the samples could be considered small; nonetheless, the focus of our study was the neuroimaging association analyses. These analyses have been conducted in a sample of hundreds of individuals, exceeding, therefore, the median sample size of neuroimaging association studies according to a recent revision (Marek et al., 2022). In this regard also, we have to point out that our structural images were obtained from two different scanners, which could represent a source of bias. Notwithstanding, we did not detect differences in neuroanatomical measures based on the two scanning sites, all the images passed the standardised quality-control protocols recommended by the ENIGMA consortium, which have been previously applied in large-scale multi-centre studies, and the scanner site was accounted as a covariate in the regressions. In terms of the genetic data, we should contemplate that our PRS estimates are pondered using SZ genetic burden and the use of other GWAS sum stats such as the corresponding to the cortical phenotypes could derive different effects. The PRS estimation method used in the present study, PRS-C+T, is the most used, and latest SZ GWAS has been conducted using the same method; however, other PRS calculation methodologies could be helpful (Ni et al., 2021). Speaking of PRS estimations, our results are based on a sample of European ancestry and SZ GWAS statistics were derived from the European cohort, then, although GWAS studies performed non-European samples converge in the same SZ’s genes and pathways (Gulsuner et al., 2020), the extrapolation of our findings to other ethnic groups should be done cautiously. Finally, data from larger samples should be highly encouraged to compare our results and replicate thereof. Also, the understanding of the role of HARs in the neurobiological roots of SZ would benefit from analyses on other brain-based phenotypes such as structural connectivity, white matter microstructure (Canales-Rodríguez et al., 2014, 2021; van den Heuvel et al., 2019), or MRI protocols related to social cognition (Fujiwara et al., 2015).

In conclusion, our study adds novel evidence on the role of the genetic variability within HARs guiding foetal neurodevelopment and shaping cortical surface area configuration in patients with SZ. The biological plausibility of our findings highlights the paramount importance of HARs in the early developmental gene regulatory machinery led us to think that these regions may contribute to bridging together the neurodevelopmental and evolutionary hypotheses in schizophrenia.

## Data Availability

The data that support the findings of this study are available from the corresponding author upon reasonable request.

## Author contributions

MG-R and MF-V conceived the study. MG-R conducted the DNA extraction and sample normalization for genotyping. EJC-R, MAG-L and PF-C pre-processed and segmented the MRI images. EP-C, PF-C, JS, JT, LT and ER-C conducted the recruitment and/or the clinical evaluation. EJC-R, RS and EP-C designed the MRI protocols. MG-R, CA and AA performed the data curation. MG-R conducted the formal statistical analyses and graphical representations with the help of CA-P, AA and ML-G. SP and MF-V were implicated in the revision of the methodology. MG-R and MF-V interpreted the results. MG-R wrote the first draft and the subsequent drafts of the paper. MF-V supervised the study activity planning and execution. MF-V and EP-C participated in the funding acquisition. All the authors reviewed and approved the final manuscript.

## Acknowledgements

The authors would like to thank the volunteers participating in the study.

